# Evaluating TESLA-G, a gamified, Telegram-delivered, quizzing platform for surgical education in medical students: a protocol for a pilot randomised controlled trial

**DOI:** 10.1101/2022.09.25.22280305

**Authors:** Matthew Song Peng Ng, Ahmad Ishqi Jabir, Tony De Rong Ng, Yi-Ian Ang, Jeng Long Chia, Darren Ngiap Hao Tan, James Lee, Dinesh Carl Junis Mahendran, Lorainne Tudor Car, Clement Luck Khng Chia

## Abstract

**Introduction:** Online multiple-choice question (MCQ) quizzes are popular in medical education due to their ease of access and ability for test-enhanced learning. However, a general lack of motivation among students often results in decreasing usage over time. We aim to address this limitation by developing Telegram Education for Surgical Learning and Application Gamified (TESLA-G), an online platform for surgical education that incorporates game elements into conventional MCQ quizzes.

**Methods and analysis:** This online, pilot randomised control trial will be conducted over two weeks. Fifty full-time undergraduate medical students will be recruited and randomised into an intervention group (TESLA-G) and an active control group (non-gamified quizzing platform) with a 1:1 allocation ratio, stratified by year of study.

We will evaluate TESLA-G in the area of endocrine surgery education. Our platform is designed based on Bloom’s taxonomy of learning domains: questions are created in blocks of 5 questions per endocrine surgery topic, with each question corresponding to one level on Bloom’s taxonomy. This structure promotes mastery while boosting student engagement and motivation. All questions are created by two board-certified general surgeons and one endocrinologist, and validated by the research team.

The feasibility and acceptability of the pilot study will be assessed by participant recruitment and retention rates, acceptability of the intervention, adherence and task completion rate, fidelity of the intervention delivery, and perception of the intervention. The effectiveness of the intervention (TESLA-G) compared to the control will be assessed by improvement in knowledge from pre- to post-intervention, learner satisfaction post-intervention, and retention of knowledge 2 weeks post-intervention.

**Ethics and dissemination:** This research is approved by Singapore Nanyang Technological University (NTU) Institutional Review Boards (Reference Number: IRB-2021-732). This study poses minimal risk to participants. Study results will be published in peer-reviewed open-access journals and presented in conference presentations.

**Trial registration number:** NCT05520671

**Strengths and limitations of this study:**

- This study contributes to the growing body of literature evaluating the use of test-based learning, messaging apps and gamification in medical education.
- The gamified, Telegram-delivered, surgical education-focused, quizzing intervention in this study will be structured in line with Bloom’s taxonomy.
- We will use quantitative and qualitative approaches to assess our intervention with the aim of informing a future randomised controlled trial.
- A potential limitation of this study is that 14 days of intervention may be insufficient to observe improvements in surgical knowledge.
- The intervention will focus on endocrine surgery and the findings may not be generalisable to other surgical or medical subspecialties.

## Introduction

### Background and rationale

Multiple-choice question (MCQ) quizzes are a well-known and widely used medium for summative assessment, especially in medical education [1,2]. Their ability to provide objective grading and immediate feedback also make them popular tools for formative assessment [3,4]. While MCQ quizzes tend to be associated with diagnostic or assessment tools, Roediger (2006) proposes that tests like these can be used to improve learning via test-enhanced learning [5]. This effect has been increasingly explored in medical education [6]. Randomised controlled trials have demonstrated how test-enhanced learning can increase acquisition and retention of new medical knowledge among medical students [7] and healthcare professionals [8]. Recent systematic review by Green (2018) also supported these findings [9].

With the progressive use of technology in medical education, online learning is becoming increasingly common. Online learning has been found to be as effective as offline learning in medical education based on meta-analyses by Pei (2019) and Vaona (2018) [10,11]. Brame (2017) suggests that making MCQ quizzes available online would allow easy access to frequent practice, which could work synergistically with test-enhanced learning to promote student learning and long-term knowledge retention [12]. This is further supported by studies that demonstrated increased exam performance after administering pre-exam online quizzes to undergraduate university students [13,14]. Kibble (2007) also established a positive relationship between unsupervised formative online quizzes and academic performance among medical students [15].

However, online MCQ quizzes are limited by a decline in quiz participation over time and an increase in drop-out rates. [13,14,16]. This was observed in several studies despite the initial enthusiasm and high take-up rate among the student body. Mitra (2015) and Johnson (2006) specifically attributed this high attrition to a general lack of motivation among the students to use the MCQ quizzes [13,16]. We aim to address this limitation of low motivation among students using online MCQ quizzes by developing Telegram Education for Surgical Learning and Application - Gamified (TESLA-G). TESLA-G is an online platform for surgical education that incorporates game elements into conventional online MCQ quizzes.

Gamification can be defined as the use of game elements such as point systems, leaderboards, and incentives in non-gaming contexts [17]. A randomised controlled trial by Barrio (2016) showed that the use of gamification in undergraduate education can boost motivation and interest [18]. A recent review by Sandrone (2021) further suggests that the use of gamification in medical education can promote engagement and motivation among learners [19]. Additionally, systematic reviews [20,21] suggest that the use of gamification for learning among healthcare professionals is as effective as other educational methods in promoting knowledge and expertise.

Nevin (2014) successfully incorporated gamification in medical education through Kaizen-IM, an online learning platform developed by the authors for internal medicine residents. The authors demonstrated that the gamification elements such as team-based score system, a leaderboard showing rankings, and badges as incentives improve medical knowledge acquisition and retention. Participants also mentioned that the game elements, especially the leaderboard, were significant motivators in their usage of the platform [22]. Notably, the use of mobile technology in gamified learning has been shown to be a motivating factor among students. Licorish (2018) found that the use of the gamified mobile platform Kahoot! among undergraduate students promoted student engagement and motivation [23]. This was attributed to students being highly proficient in mobile technology, which greatly contributed to significant student enjoyment in trying out applications and games that have been built specifically for mobile platforms [23].

Instant messaging applications have been used as a supplementary tool to medical education, [24], and several studies have shown an improvement in knowledge level through the use of these platforms [25–28], most notably with the application WhatsApp. More recently however, the increasingly popular messaging application Telegram has been trialled for use in medical education in the context of the COVID-19 pandemic [29,30]. Telegram was the most downloaded app in the world in January 2021 [31], and still remains as the top five most popular messaging apps globally in 2022 [32]. Not only is this app accessible on almost all computer and mobile platforms, the increasing use of Telegram in the upcoming years would mean that it is very likely that participants will already have this app installed in their devices even before the commencement of the study. By tapping on existing Telegram features and its well-documented Application Programming Interface (API), we will implement TESLA-G to provide flexible and convenient surgical education anytime and anywhere. Hence, our online, gamified quizzing platform TESLA-G will be delivered using Telegram with the aim of easier delivery of the intervention and greater uptake. We intend to evaluate our intervention in a stepwise manner in line with the UK Medical Research Council’s guidance for developing and evaluating complex interventions [33]. In this pilot study, we will evaluate the feasibility and acceptability of delivering our intervention with the aim of informing a future randomised controlled trial.

### Objectives

The main objective of the pilot randomised controlled trial is to evaluate the feasibility and acceptability of a gamified, online, Telegram-delivered quizzing platform TESLA-G compared to conventional MCQ quizzes for surgical education among medical students.

More specifically, we will investigate:

1. The feasibility of recruitment, specifically the duration required to complete the recruitment process, the recruitment strategy, randomisation and stratification strategy, and participants’ retention rate throughout the proposed intervention period.
2. The acceptability of the intervention to medical students, in terms of its technical, pedagogical, and educational content (surgical content).
3. The participants’ adherence to the intervention, in terms of the number of completed quiz questions, frequency of use, and average daily quiz completion rate.
4. The fidelity of the intervention protocol, in terms of whether the assessment materials, technical implementation of the intervention, and study procedures are delivered and successful.
5. The participants’ experience of the intervention, by inviting a purposive sample of the medical students to share their views via semi-structured interviews after the intervention.

Our secondary objectives are:

1. To evaluate the effectiveness of TESLA-G compared to conventional MCQ quizzes in improving surgical knowledge by comparing the change in scores between the pre- and post-intervention tests.
2. To compare students’ satisfaction with TESLA-G compared to conventional MCQ quizzes using a post-intervention satisfaction survey.
3. To evaluate the effectiveness of TESLA-G compared to conventional MCQ quizzes in retention of surgical knowledge which will be determined by comparing the scores a follow-up knowledge test administered 2 weeks after the post-intervention test

## Methods and analysis

### Trial design

We report this protocol in line with the SPIRIT (Standard Protocol Items for Randomised Trials) recommendations [34]. This is an online, pilot randomised controlled trial with two parallel active groups. It will involve an intervention group and an active control group. Participants will be randomised into these two groups with a 1:1 allocation ratio stratified by year of study.

### Study setting

The study is an online study conducted on Telegram. The study will recruit first to fifth year medical students from a medical school in Singapore.

### Eligibility criteria

Eligible participants must be

1. at least 18 years of age;
2. currently enrolled in a full-time 5-year undergraduate programme in the medical school that leads to the Bachelor of Medicine and Bachelor of Surgery (MBBS);
3. willing and able to provide consent for participating in the entire duration of the study including all pre- and post-study assessments.

### The intervention

TESLA-G is a novel gamified quizzing platform aimed at improving surgical knowledge among undergraduate medical students. For the purpose of this study, we will evaluate the use of TESLA-G in the learning of endocrine surgery among medical students. Our platform is designed based on Bloom’s taxonomy of learning domains, which is a widely applied and researched framework to construct learning objectives [35]. Questions will be created in blocks, where each block will test a specific topic in endocrine surgery. Each block has 5 questions, and each question corresponds to the first five levels of the Bloom’s taxonomy (Remember, Understand, Apply, Analyse, Evaluate) and each level in game (Table 1). The questions are structured in this way to promote mastery in endocrine surgery while boosting student engagement and motivation.

**Table 1:** sample block of 5 questions corresponding to different Bloom’s taxonomy levels

|  |  |  |  |
| --- | --- | --- | --- |
| Level 1 | Remember | Questions here encourage students to recognise and recall facts. | FNAC thyroid reveals a benign follicular nodule. What Bethesda category would this be in? |
| Level 2 | Understand | Questions here motivate students to understand the meaning behind the memorised facts. | FNAC thyroid reveals a Bethesda category II follicular nodule. What is the probability of malignancy? |
| Level 3 | Apply | Questions here allow students to apply their knowledge in a clinical setting. | 31 year old lady was referred by a GP who noticed a goitre incidentally. Which of the following clinical features would be most suspicious of malignancy? |
| Level 4 | Analyse | Questions here would expect students to analyse the clinical presentation and use logical deduction to figure out the differential diagnoses. | 31 year old lady was referred by a GP who noticed a goitre incidentally. Further workup shows a solitary nodule in the right thyroid 2.5cm. FNAC reveals Bethesda category II follicular nodule. Patient thyroid function is normal. |
|  |  |  | What is the most likely differential? |
| Level 5 | Evaluate | Based on a constellation of clinical findings, students are expected to critically examine and select the most appropriate investigations or management options. | 31 year old lady was referred by a GP who noticed a goitre incidentally. Further workup shows a solitary nodule in the right thyroid 2.5cm. FNAC reveals Bethesda category II follicular nodule. Patient thyroid function is normal.<br>What is the most appropriate management? |

For this study, we aim to create 56 blocks of 5 questions, totalling 280 questions. All questions will be created by two board-certified general surgeons and one endocrinologist, and validated by the research team.

The aim of the game is for players to get as many points as they can before the timer runs out. Each game will feature only one block of 5 questions, allowing students to focus on a specific topic in endocrine surgery per game (Fig 1).

**Fig 1:**
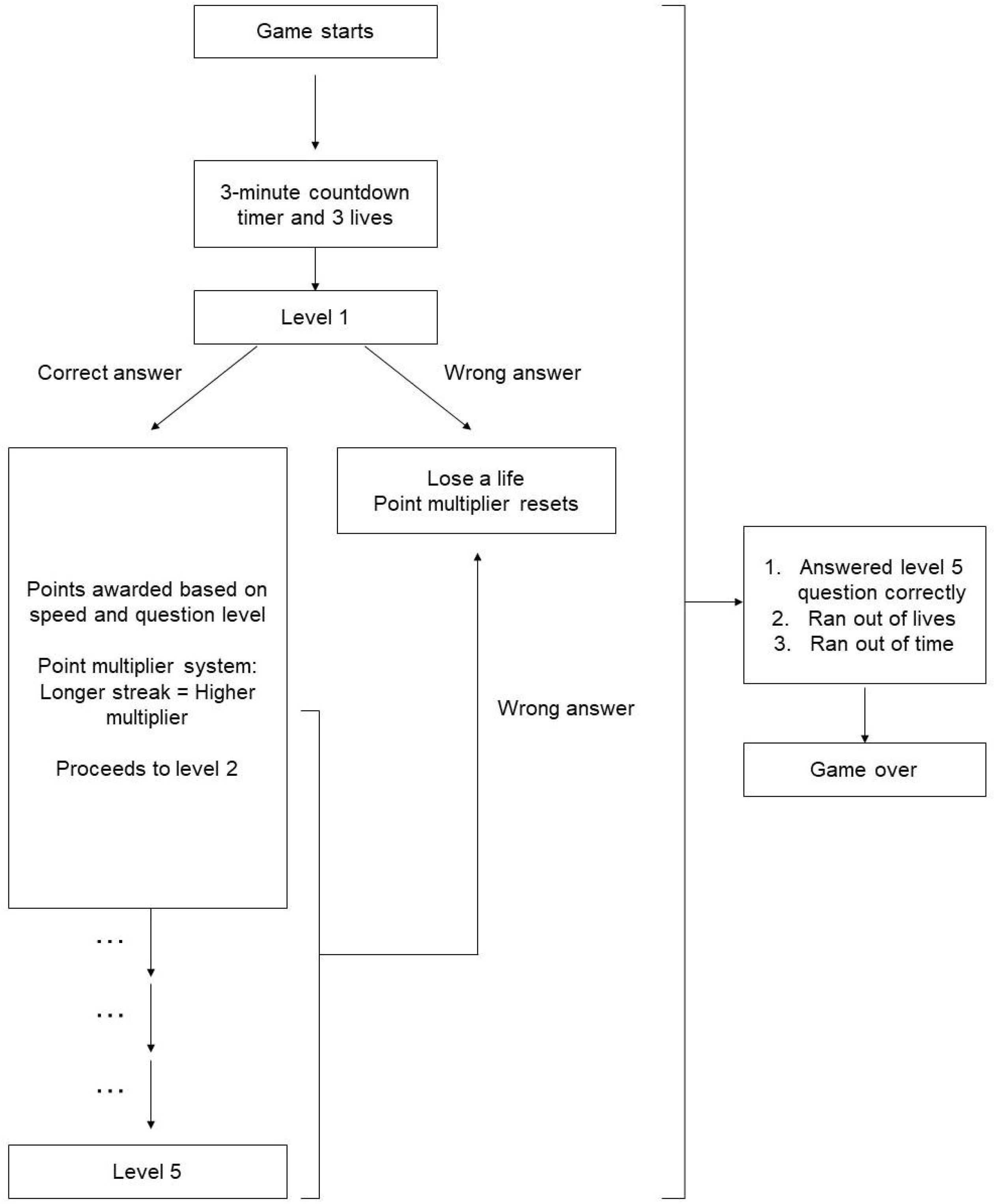
flowchart showing the run-through of the game

Below is a run-through of the game:

1. The game starts at Level 1 with a 3-minute countdown timer and 3 lives.
2. For every question answered correctly, students will be awarded points based on their speed and the question level. An explanation of the question will also be shown. They will then progress to the next level up till level 5.
3. Consecutive correct answers will be rewarded via a point multiplier system. The longer their “streak” of correct answers, the higher the multiplier. It should be noted that levelling up does not require consecutive correct answers.
4. If they get a question wrong, they lose one life, and the point multiplier system resets. They must then answer that question again until they get it correct.
5. The game ends if the level 5 question is answered correctly, if all 3 lives have been used, or if the timer runs out. The total score is then tabulated.

To ensure proper mastery of surgical concepts, the countdown timer will pause in between questions for students to refer to the explanation pop-up. Students will be given as much time as they need to read the explanation and understand the information presented before they move on to the next question. There is no score deduction for spending too much time reading the explanation after each question.

To promote competition-based learning, all students are ranked against each other based on their points, and a leaderboard that displays the top 10 students is sent to all students daily. We also allow students to track their own learning by providing a personalised dashboard for students to see their score history and progress. Finally, to promote consistent usage of TESLA-G, a bonus point multiplier is awarded for every consecutive day the game is played.

### Timeline

The study flow is summarised in Fig 2 and the logic model is shown in Fig 3. Before any participants are recruited, all tests and surveys will first be piloted by all researchers involved in this study. Qualitative feedback will be independently obtained from at least 3 researchers, and tests and surveys will be amended accordingly.

**Fig 2:**
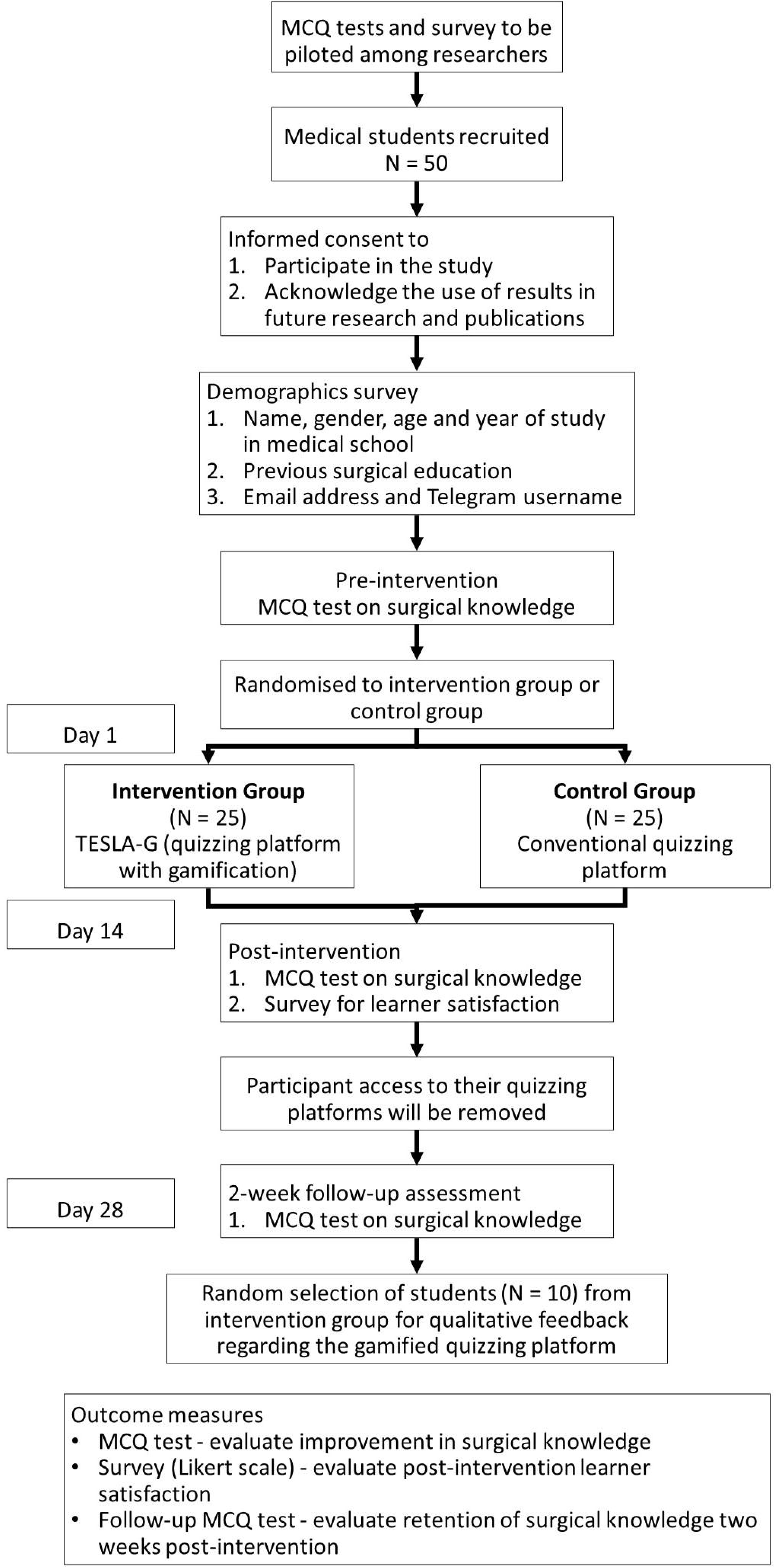
study flow diagram

**Fig 3:**
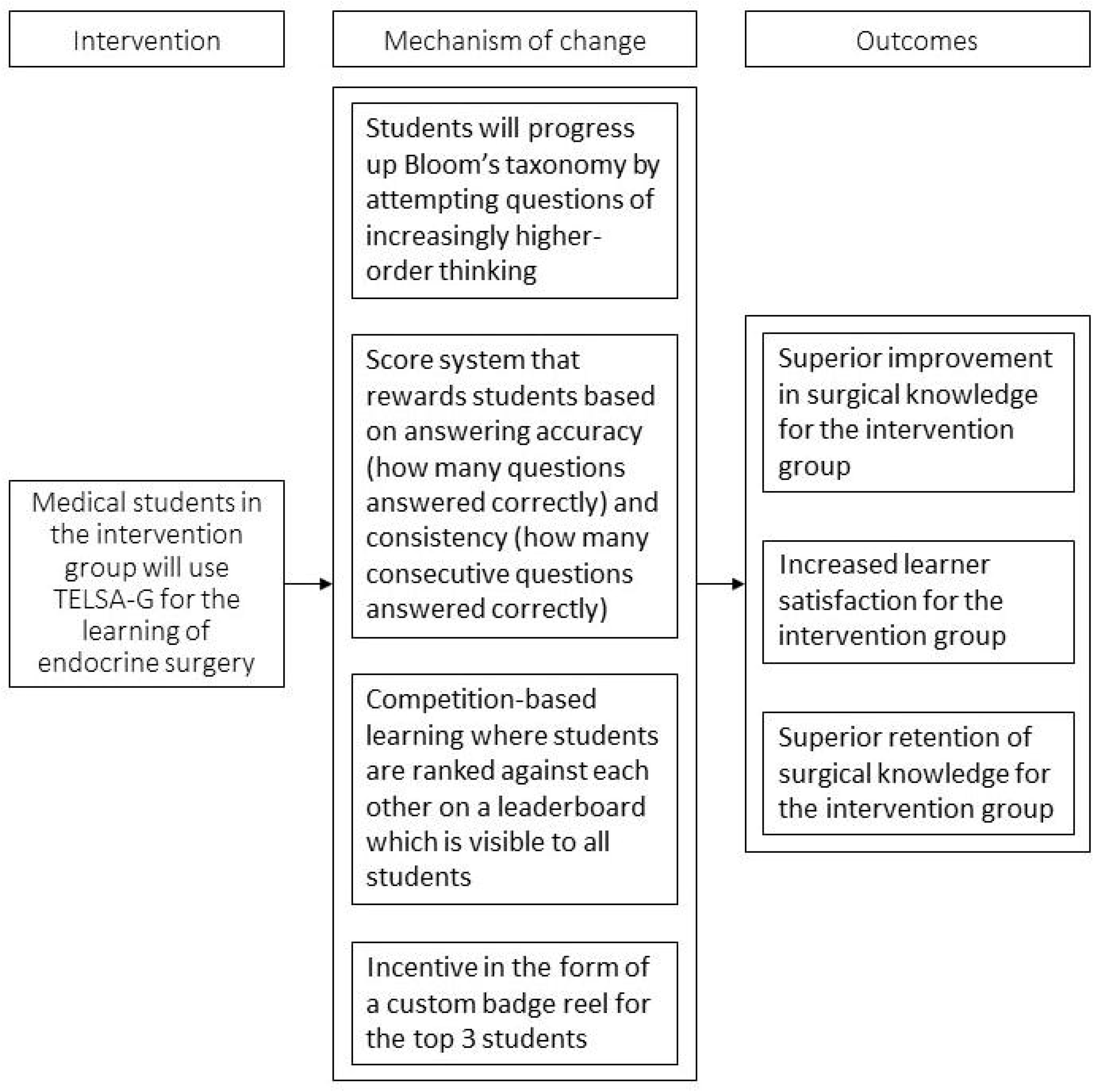
logic model of the study

#### (i) Pre-intervention

Fifty medical students (*N* = 50) will be recruited for the study based on the eligibility criteria. After obtaining informed consent, all participants will be expected to complete a demographics survey. Basic information about name, gender, age and year of study in medical school will be collected.

The Telegram username and email address of each participant will also be collected and verified. This is for the main purpose of disseminating information regarding our study. These include important deadlines to take note of, instructions on how to access the quizzing platforms and links to the other surveys required for our study.

All participants will then complete a pre-intervention knowledge test comprising 20 MCQs on endocrine surgery over a 30-minute period. This will determine the baseline surgical knowledge level among all participants.

#### (ii) Intervention

Participants will be verified to have completed all the above before being randomised into either the intervention group or the control group. Participants will then be provided with detailed instructions on using either TESLA-G for the intervention group or the conventional quizzing platform for the control group.

Participants in the intervention group will be provided access to TESLA-G as previously described. A link to access TESLA-G will be sent to participants from an automated Telegram bot; this access will be provided for 14 days. Participants in the control group will be given access to a conventional quizzing platform, which will be a non-gamified version of TESLA-G. Whenever a participant enters the platform, a question stem and five options will be displayed. The participant will select an option, and the correct answer appears with the explanation provided. The platform will then send the next question, and the process continues, until the participant decides to exit the platform or all the questions in the platform have been answered. Similar to the intervention group, the questions will be queued in blocks where each block corresponds to a specific topic in endocrine surgery. Within each block, questions will be randomised, that is, they will not be queued based on increasing levels of Bloom’s taxonomy. Participants will also not be notified what level of Bloom’s taxonomy each question is classified under. Just like the intervention group, participants will be sent the link to access this platform from an automated Telegram bot, and they will be able to use the platform for 14 days.

For both the intervention and the control groups, all questions will be made available to all participants from the beginning of the study. If any participant does not use their quizzing platform for more than 48 hours, a Telegram message will be sent as a gentle reminder to continue participation. This would promote continuous usage of the quizzing platforms throughout the entire duration of the study.

Of note, throughout the usage of either quizzing platform, participants will be shown new and previously completed questions, for the purpose of mastering new knowledge and reinforcing past concepts, respectively.

Participants will use their assigned platform at their own pace and time. During this period, user metrics will be collected from both the intervention and the control groups including overall platform, per-participant and per question metrics as presented in Table 2.

**Table 2:** game metrics to be collected from both groups. A block refers to a question block of 5 questions, as previously described.

| Overall platform metrics | Per-participant metrics | Per-question/block metrics |
| --- | --- | --- |
| Game performance<br>• Number of logins per hour, per day and over the full study duration<br>• Total usage duration per hour, per day and over the full study duration | Participant engagement<br>• Number of daily logins<br>• Number of questions and blocks that were completed daily<br>• Number of questions and blocks that were re-attempted daily<br>• Time of logins<br>Participant performance<br>• Daily usage duration<br>• Individual scores per gameplay | Question engagement<br>• Number of users who completed each question and block<br>• Amount of time spent on the explanation pop-up<br>Question performance<br>• Distribution of answers for each question<br>• Number of users who obtained the correct answer for each question<br>• Numbers of users who obtained a full completion for each block |

Finally, throughout the duration of the intervention, participants will be able to contact a designated study team member via Telegram or via email for any assistance with potential technical difficulties with using their quizzing platform.

#### (iii) Post-intervention

After 14 days, all participants will complete a post-intervention knowledge test which again comprises 20 MCQs on endocrine surgery over a 30-minute period. This test will be of a similar standard to the pre-intervention test as described earlier. Improvement in surgical knowledge will be measured as the difference in knowledge test scores pre-intervention and post-intervention.

All participants will also complete a post-intervention learner satisfaction survey. This will be in the form of a Likert scale adapted from the System Usability Survey (SUS) [36] and the Student Evaluation of Educational Quality (SEEQ) Questionnaire [37]. The survey will find out if the intervention group of students are more satisfied than the control group. To incentivise participants, the top three participants of the leaderboard will receive small tokens of appreciation in the form of a certificate and a custom badge reel with the TESLA-G logo.

After another 14 days, participants will be asked to do a follow-up knowledge test which once again comprises 20 MCQs on endocrine surgery over a 30-minute period. At this point, participants in both the control and the intervention groups will not have access to their respective online quizzing platforms. This test will be of a similar standard to the post-intervention test as described earlier. Any retention in surgical knowledge will be measured as the difference in knowledge test scores post-intervention and follow-up.

After the follow-up knowledge test, ten participants from the intervention group will be purposefully selected for individual interviews. Two participants - one who has completed at least 80% of the quizzes and one who has not - will be selected from each of the five academic years of the medical school. We aim to obtain qualitative feedback regarding the overall experience of TESLA-G, along with the benefits, drawbacks, receptiveness and usefulness of the platform as a supplement to surgical education. The interviews will be conducted based on the interview guide (Supplementary File, Interview Guide) which has been adapted from a previous exploratory study on using Telegram for surgical education [38]. The interviews will also be piloted on up to five students and/or research team members prior to the beginning of the study.

The interviews will be conducted individually, either online or in-person, and will last 60 minutes. Written and verbal consent will be taken, before the interviews are recorded to be transcribed into text data for qualitative analysis. The audio recordings from the interviews will be transcribed by an automated software and proof-read to ensure that the transcripts are accurate. The completed transcripts will not contain any identifiers.

### Outcome measures

A mixed-method approach will be used to establish the feasibility and acceptability of the intervention as primary outcomes. Our secondary outcomes will be improvement of surgical knowledge between the control and intervention group and potential adverse effects. Finally, qualitative feedback from the participants regarding their experience of the intervention will be thematically analysed.

#### (i) Feasibility

The feasibility of the intervention will be assessed quantitatively as shown below. These are the goals for our intervention:

- Enrollment of 50 participants in a month
- Retention of at least 75% of participants who are enrolled
- Completion of at least 80% of the quizzes

Success in achieving all three goals will indicate that it is definitely feasible to conduct a full-scale randomised controlled trial (RCT) while achieving two out of three goals will indicate that it is probably feasible. Achieving less than two goals will suggest that a full-scale RCT is not feasible with the current procedure.

In addition to the above outcomes, we will also determine:

- The feasibility of the randomisation procedure to ensure equal number of participants within the different university years in each stratum within the intervention and control groups
- Amount of time on app and on task
- Feasibility of the delivery method for the pre-, post- and follow-up assessments
- Number of app crashes
- Number of app exits during task

#### (ii) Acceptability

The acceptability of the intervention and the study procedure will be assessed both quantitatively and qualitatively. Quantitative data will be collected via a post-intervention learner satisfaction survey for both the control and intervention groups. This survey will consist of two parts - a system satisfaction questionnaire and a content satisfaction questionnaire.

The system satisfaction questionnaire measures the system acceptability score via a System Usability Survey (SUS) [36]. It is a 10-item questionnaire with a 5-point Likert scale that ranges from 1 (strongly disagree) to 5 (strongly agree). This widely used usability scale will be used to compare the relative usability between TESLA-G and our conventional quizzing platform based on a normalised score. For the SUS, we aim to get an average score of at least 70 which would indicate Grade B based on the Sauro and Lewis (2016) curved grading scale [39].

The content satisfaction questionnaire measures the content satisfaction score and is loosely adapted from the Student Evaluation of Educational Quality (SEEQ) Questionnaire [37]. The SEEQ evaluates 9 distinct components of teaching effectiveness with a 5-point Likert scale that ranges from 1 (very poor) to 5 (very good). This validated study has been rigorously evaluated in higher education [40,41] and has been used in clinical education [42,43]. This questionnaire will compare the relative benefit of content delivery between TESLA-G and our conventional quizzing platform. For this, we aim for a mean score of at least 6.0 for TESLA-G with a significantly higher score when compared to the conventional quizzing platform.

#### (iii) Improvement of surgical knowledge

The difference in the improvement of surgical knowledge between the control and intervention group will be determined by comparing the scores of pre- and post-intervention knowledge tests. These tests consist of questions on endocrine surgery which will be created separately by two board-certified general surgeons and one endocrinologist. All questions will also be validated by the research team. It should be noted however, that the study will not be sufficiently powered to identify the comparative effectiveness between the control and intervention groups in terms of improvement of surgical knowledge overtime. Hence the analysis will be primarily conducted to determine any potential adverse effects and increase in the surgical knowledge within each group, and secondarily between groups. This will be done using a confidence interval of 95% using a small effect size of 0.2 [44].

#### (iv) Qualitative feedback

The interview transcripts obtained from the participants who used TESLA-G will be coded into various categories, based on the overall question about how TESLA-G has been useful in improving surgical knowledge. Recurring themes will then be identified and substantiated with illustrative quotes. These themes as well as verbatim quotes will then be incorporated into the discussion of the findings chapter as with all qualitative studies.

### Sample size and recruitment

Fifty participants (*N* = 50) will be recruited, i.e. 25 participants per arm. This was based on a study by Whitehead *et al*. (2015) which suggested that 25 participants for each arm is optimum for studies with small effect size (between 0.1 to 0.3) for 90% power [45]. Additionally, a purposive sample of at least ten medical students (*N* = 10) will be invited to share their views of the intervention via semi-structured interviews. This is based on consideration of the resources available.

Our study advertisement will be disseminated to all medical students from LKC School of Medicine via the respective Telegram group channels for each cohort. We will also advertise through advertisement posters and recruitment calls during regular lectures/seminars as well as through personal contacts.The study advertisement has a link to a secure registration form hosted by the research IT department. Every participant will be expected to fill in this registration form. This registration form will collect the name and email address of each participant. A letter of informed consent will be emailed to each participant, and he/she will be expected to provide consent in order to be considered as recruited for the study.

### Blinding and randomisation

Participants will be partially blinded. They will not know if they have been given the conventional quizzing platform or TESLA-G. Instead, they will be told that they have been randomly allocated to one of two different quizzing platforms. All researchers involved in statistical analysis will be blinded as far as possible. They will not know which group is the control and which group is the intervention until the analysis is complete.

Only one researcher will be involved in dissemination of information to participants, and s/he will not be blinded. This is necessary because both quizzing platforms will have different instructions on accessing and using the platforms. It is important that the correct instructions are sent to each participant. This researcher will also be involved in answering any platform-related queries from participants throughout the study.

Participants will be stratified by their year of study. Following this, participants will be randomly allocated into either the intervention or the control group. Permuted block randomisation will be conducted for each stratum using a computerised random number generator to ensure a 1:1 allocation ratio and equal group sizes. To ensure allocation concealment, this randomisation process will be conducted and kept confidential by a trusted individual outside of the research team.

### Statistical analysis

Analysis of quantitative data comparing TESLA-G with conventional quizzing platforms will be performed using commercial statistical software (SPSS for Windows, version 22.0, Chicago, Illinois, USA). All categorical variables will be described as percentages and compared by Chi-squared test. The primary outcomes will be analysed descriptively with outcomes being the means, standard deviations, and interval estimates of variables relating to the feasibility and acceptability of the study. The secondary outcomes relating to the effectiveness of TESLA-G, such as the knowledge test scores and learner satisfaction scores for each group, will be presented using descriptive statistics as well. Comparison of scores will be done with paired t-test analysis and repeated-measures analysis of variance (ANOVA) statistics using a confidence interval of 95% [44].

Analysis of the qualitative feedback from the interviews will be performed with a parallel template analysis of inductive and deductive analysis until saturation is reached. Open coding of the transcripts will be performed by at least two independent coders. Qualitative data will then be thematically analysed through a selected qualitative data analysis software. The codes identified will subsequently be categorised into themes. Number and frequency of responses will also be tabulated.

### Data monitoring and harms

Participants will be encouraged to comply with the protocol as far as possible. Researchers will check that participants meet the submission deadlines of all pre- and post-intervention tests and surveys. Researchers will also ensure that all tests and surveys are properly filled up.

Participants who do not comply with the study protocol (such as submitting tests late or submitting surveys with all items answered with the same response) will not be removed from the study as analysis will be done with an intention-to-treat approach.

This is a very low-risk study. Throughout this study, students will be encouraged to inform the research team and their university’s student support services if they experience any issues, harm or psychological distress. Such incidents will also be recorded and reviewed for future improvement. A data monitoring committee (DMC) will not be needed.

### Patient and public involvement statement

Patients or members of the public will not be involved in the design, conduct, reporting or dissemination plans of this research.

## Discussion

### Implications

The primary objective of this pilot trial is to provide important information regarding the feasibility and acceptability of TESLA-G among medical students, with the aim of informing a future full-scale RCT. As a secondary objective, a demonstration of statistically significant improvements - in surgical knowledge, learner satisfaction and knowledge retention - by TESLA-G in this pilot trial may suggest a greater effect in a full-scale RCT with a larger sample size.

We aim to contribute to the growing body of literature evaluating the use of test-based learning, messaging apps and gamification in medical education. One key strength of TESLA-G is that it is structured in line with Bloom’s taxonomy [35], which has been recently applied in clinical learning, such as in clinical simulation [46] and in clinical-oriented surgical education [47]. Medical students in our study will attempt blocks of 5 questions that are specifically designed according to the increasing difficulty levels of Bloom’s taxonomy.

### Limitations

Firstly, the effectiveness of participant blinding is limited due to the difficulty in preventing communication among participants. It would be unfeasible to physically or digitally isolate the intervention and control groups from each other for the entire study duration. Participants will however be blinded as to whether their online quizzing platform belongs to the intervention group or the control group, and constant reminders will be given to participants before and during the study to ensure they do not communicate with other study participants or compare the test and control platforms.

Secondly, the intervention duration of 14 days may be too short a time to allow the beneficial effects of TESLA-G (e.g. improvement in surgical knowledge) to manifest. If this is so, a full-scale RCT with a larger sample size may demonstrate a greater effect.

Lastly, endocrine surgery is the only surgical topic evaluated in the study, hence results showing the effectiveness of TESLA-G may not be generalisable to other surgical subspecialties, or other medical specialties. Future work would involve expanding TESLA-G to encompass other surgical subspecialties, and using TESLA-G as a template to develop more gamified online platforms that would better cater to the other medical specialties.

## Supporting information

Reporting checklist for protocol of a clinical trial

## Data Availability

All data produced in the present study are available upon reasonable request to the authors

## Ethics and dissemination

### Research ethics approval

This research has been approved by NTU Institutional Review Boards (Reference Number: IRB-2021-732).

### Consent

All participants will be expected to read and sign a letter of informed consent before they are considered as recruited into the study. For participants below the age of 21, this letter will be co-signed with their parents. This letter would include the study information, participant rights and contact details of the Project Investigators.

Participants will be reminded that they can withdraw from the study at any point in time without giving any reasons by informing the principal investigator and all data collected of the participant will be discarded.

### Data management and confidentiality

All data collected during the study will be kept confidential. Any identifiable information will only be stored on the university’s secure storage folders. Only the principal investigators will have access to these data for the sole purposes of verification and participant follow-up, if needed. Data stored in Telegram servers and TESLA-G servers will be indexed according to the participants’ computer-generated unique ID to maintain anonymity, and at no point in time will these servers store identifiable information from any participant. At the end of the study, only anonymised data indexed by unique IDs will be made available by the principal investigators to the research team for data analysis.

Data will be backed-up regularly. Personal data will never be used in a publication or presentation. All data collected will be kept in accordance with the University’s Research Data Management Policy. Research data used in any publication will be kept for a minimum of 10 years before being discarded.

### Dissemination plan

Study results will be published in peer-reviewed open-access journals and presented in conference presentations.

## Authors’ contributions

CLKC and LTC conceived the study concept. MSPN, AIJ, YIA and TDRN drafted the manuscript and obtained funding. MSPN developed the study design and statistical analysis plan. JLC and DNHT developed the software aspects of TESLA-G. CLKC, LTC and AIJ provided critical revision. All contributors have approved the final version of the manuscript.

