## Supplementary File for "Evaluating TESLA-G, a gamified, Telegram-delivered, quizzing platform for surgical education in medical students: a protocol for a pilot randomised controlled trial"

**Interview Guide**

Questions on demographics

| No. | Questions |
| --- | --- |
| 1 | Which year of medical school are you currently in? |
| 2 | Have you completed your general surgery rotation? |
| 3 | Are you currently taking part in your general surgery rotation? |
| 4 | How long have you been using Telegram? |
| 5 | What do you primarily use Telegram for? |
| 6 | What other messaging apps do you use? |

Questions for TESLA-G

| No. | Questions | Prompts |
| --- | --- | --- |
| 1 | What role has mobile learning (i.e., learning delivered via mobile devices) had on your education? | During this pandemic; In general; In surgical education; In other areas; Types of mobile learning used; Pros/Cons to its use |
| 2 | What is your view on the use of messaging apps (e.g. Telegram or WhatsApp) in your medical education? | Areas in which they can be used; Pros/cons of using them; Preferences |
| 3 | What is your view of TESLA-G? | Benefits/limitations |
| 4 | What impact did the TESLA-G have on your surgical competency? | Knowledge; Skills |
| 5 | What impact did TESLA-G have on your attitudes towards surgery? | Surgical training; Surgery as a speciality/choice of residency |
| 6 | What is your view of the potential inclusion of TESLA-G in formal undergraduate surgical education in future? | Benefits/limitations |
| 7 | How can TESLA-G be improved? | Additional content; Features |
| 8 | What is your view on other similar learning platforms that you have used? | Similarities/differences; Advantages/disadvantages |
| 9 | What is your view on the use of such learning platforms in other areas of your medical education? | Areas; Format of the channel; Advantages; Disadvantages |
| 10 | Are there any other observations or comments that you would like to share with me today? |  |
